# Postpartum suicidal ideation in Austria and Germany during the COVID-19 pandemic

**DOI:** 10.1101/2024.03.15.24304383

**Authors:** C. Florea, J. Preiß, M. Angerer, M. Schabus

## Abstract

**Introduction:** Postpartum suicidal ideation is a significant concern, as it poses a risk for future suicide attempts, particularly in high income countries, where suicide ranks among the leading causes of death for postpartum mothers. The literature indicates a global average prevalence of postpartum suicidal ideation of approximately 7%, but for Austria and Germany there are few studies on this subject.

**Methods:** In a web-based survey for Austrian and German mothers of children born during the COVID-19 pandemic, several measures of mental health (depression, stress), social support and other parenting and pandemic-related questions were assessed in 1964 mothers. Based on the answers for the last item of the Edinburgh Postpartum Depression Scale, the suicidality risk and the presence or absence of suicidal ideation were computed. Furthermore, possible risk or protective factors for suicidality were investigated.

**Results:** The prevalence of suicidal ideation was 7.3%, which is in the range of the global prevalence reported in the literature, but two times higher than previous reports on German mothers. The three strongest risk factors for suicidal ideation were (i) high levels of stress (increased risk by 350%), (ii) a lack of perceived social support (increased risk by 265%), and (iii) a perceived negative effect of the pandemic on the relationship with the partner (increased risk by 223%). Not receiving help from family and friends, having a lower income, and feeling negatively impacted by the pandemic also significantly increased the risk of suicidal ideation.

**Discussion and conclusion:** The results indicate a higher prevalence of suicidality than previously reported in German mothers, and confirm the risk factors previously associated with depression and suicidality. These risk and protective factors could be targets of social and public health policies, while the first step should be a general screening program for suicidality in this population group.

## Introduction

### Postpartum depression

Depression is a mood disorder that manifests through a lack of interest and joy in most activities of daily life, which persists for at least two weeks. During the postpartum period, depression not only affects the mother, but also her family, including the partner (Shorey et al., 2018), and the especially vulnerable infant (Leiferman, 2002). Postpartum depression is a global public health issue, with a prevalence of about 17% in healthy mothers (Shorey et al., 2018), though strongly varying from country to country (Halbreich & Karkun, 2006). Studies on German and Austrian populations also found varying prevalence of postpartum depression symptoms: 5% with an Edinburgh Postnatal Depression Scale (EPDS) cut-off of 13 points (Figueiredo et al., 2004), 17.1% with a cut-off 9.5 points (Bergant et al., 1998; v. Ballestrem et al., 2005, as cited by Halbreich (Halbreich & Karkun, 2006) or 42% with an EPDS cut-off of 9.5 in an overview study of our lab (Florea et al., 2023). The differences in the prevalence values might, on the one hand, stem from a higher EPDS cut-off point in the study of Figureiredo and colleagues (2004), in which a score of ≥ 13 was considered as depression. This raises the specificity of the EPDS test to 95%, but at the same time lowers its sensitivity to just 66%, meaning that it misses 33% of depression cases (Levis et al., 2020). On the other hand, the studies might differ in their timing, with the studies of Figureirerdo et al. (2004), Bergant et al. (1998) and Ballestrem et al. (2005) being performed before the COVID-19 pandemic (i.e., in 1998 and 2004), while the study of our lab (Florea et al., 2023) took place during the pandemic when depression levels increased (Ceulemans et al., 2020; Zanardo et al., 2020).

### Suicidal ideation

One of the symptoms of depression is the presence of thoughts of self-harm and suicide (American Psychiatric Association, 2013). Suicidality can arise in the absence of depression, but often they overlap (Garman et al., 2019; Iliadis et al., 2018), so that scales that identify both have been developed. The EPDS (Cox et al., 1987) is one of the most often used tools for the screening of postpartum depression and suicidal ideation (Xiao et al., 2022). It is a 10-item scale that evaluates the presence of depression symptoms over the last seven days and contains an item inquiring about thoughts of self-harm or suicide. A positive score on this item (indicating that the mother has thoughts of self-harm or suicide) should lead to immediate discussion with a health care specialist, so that any appropriate referrals can be made and any harm for the mother or the baby can be avoided.

Thoughts of suicide and self-harm are an indicator for future suicidal attempts (Orsolini et al., 2016), and during the postpartum period, suicide is one of the leading causes of maternal death in high-income countries (Oates, 2003). The prevalence of postpartum suicidal ideation is around 7%, and the highest rates have been found at around 4-months after delivery (Xiao et al., 2022). Factors that have been associated with suicidal ideation include younger age, higher parity (Howard et al., 2011), unpartnered relationship status, experiencing partner violence or abuse (Orsolini et al., 2016), lower income group (Begum et al., 2021), or obstetric and neonatal complications (Ayre et al., 2019). Furthermore, in an extreme situation such as during the COVID-19 pandemic, the prevalence of thoughts of self-harm might increase compared to the time prior to the pandemic (Wu et al., 2020).

### Postpartum suicidality risk in Austrian and German mothers

There are few studies on postpartum suicidality risk in Austrian and German mothers (Martini et al., 2019), despite this group being at high risk for depression due to the postpartum hormonal changes, changes in sleep schedule, and general challenges of taking care of an infant. In a previous study from our lab on Austrian and German mothers of children born during the pandemic, we found high rates (42%) of depressive symptoms during the COVID-19 pandemic, and the main factors that influenced the depression scores were the social support, the perceived negative influence of the pandemic, and the presence of an effective coping mechanism (Florea et al., 2023).

In the current report, we investigated the sample of our previous study (Florea et al., 2023) in more detail. Specifically, we focused on the prevalence and risk factors of suicidal ideation in postpartum mothers during the COVID-19 pandemic. Based on the previously published results, which indicate that social support has a protective influence against depression, we hypothesize that it has the same protective effect on the suicidal ideation. Negative effects of the pandemic were correlated with higher depression scores in the same population, which leads us to the hypothesis that they also correlate with a higher risk of suicidal ideation. Furthermore, we hypothesize that the risk of suicidal ideation is higher in case of lower income, lower education levels, worse access to a midwife, or high stress levels. Further, we explore what other factors might correlate with an increased risk of suicidal ideation.

## Methods

The data was collected via an online survey published in Austria and Germany between 18.05.2021 and 01.07.2021, a period during which restrictions were in place to prevent the spreading of the coronavirus. Parents and other caregivers of children born since the first lockdown started (16.03.2020), aged between 18 and 65 years of age and living in Austria or Germany were eligible for the survey. Only complete datasets were analyzed, and for the current report only the biological mothers of the children were selected. For more details about the study design, participants, and survey content, see Florea et al., 2023.

The survey included questions specific to the pandemic situation, to the pregnancy, birth and postpartum period, and scales to measure stress (Perceived Stress Scale [PSS], Cohen et al., 1983; E. E. Schneider et al., 2020) and depression (Edinburgh Postnatal Depression Scale [EPDS], Bergant et al., 1998; Cox et al., 1987). Suicidal ideation is defined here as a positive answer (a score > 0 on a scale from 0 to 3) to the item 10 of the EPDS. The item questions the participant on any thoughts of self-harm and their frequency over the last seven days: never, seldom, sometimes, or often. From this question, we extracted two variables: (1) the suicidal ideation as a binary variable: 0 if the mothers report to never have suicidal thoughts, and 1 if they do, irrespective of frequency, and (2) the suicidality score, quantifying how often the person has suicidal thoughts, on a scale from 0 (never) to 3 (often). The (1) suicidal ideation is the variable we use to compute the relative risk ratio between the groups, since it is binary and therefore can be counted as an indicator of incidence, which makes it better suited for the calculation of relative risk. The (2) suicidality score is the variable we use to see if groups differ significantly, since it is an interval scaled variable and contains more information.

To see how different factors influence the suicidality score and the incidence of suicidal ideation, we split the participants in two or three groups based on the respective grouping variable, for example, low vs. high social support (by median split), or negative change/no change/positive change in the relationship with the partner during the pandemic. To identify possible differences between groups, we (1) compared the suicidality score between the two/three groups using the Wilcoxon rank sum test or Kruskal-Wallis test. Afterwards, we (2) calculated the percentage of suicidal ideation in each group and calculated the relative risk by comparing the percentages between groups. In case there were three groups, we compared each of the extremes (e.g., negative/positive change, or pre/post-term birth) with the middle (e.g., no change, or at-term birth). In case there were two groups, we compared them with each other. The relative risk ratio was computed using the following formula (Noordzij et al., 2017):

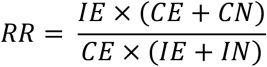

Where *RR* is the relative risk of having suicidal thoughts in the “exposed” (at-risk) group, *I* is the “intervention” group (“exposed group”), *C* is the “control” group, *E* means “event” (i.e., suicidal ideation), and *N* means “non-event” (i.e., no suicidal ideation). For a practical example, consider the low and high social support groups: low social support is the “intervention”, high social support is the “control”, because we want to see if low social support adds a risk of suicidal ideation compared to high social support. The event is suicidal ideation. Therefore, four groups emerge: low social support with suicidal ideation (*IE*), low social support without suicidal ideation (*IN*), high social support with suicidal ideation (*CE*) and high social support without suicidal ideation (*CN*) (Table 1). The relative risk ratio of suicidal ideation for mothers with low social support compared to mothers with high social support is, practically, the probability of suicidal ideation in the group of low social support *IE*/(*IE* + *IN*) divided by the probability of suicidal ideation in the group of high social support *CE*/(*CE* + *CN*). A relative risk of 1 means no difference between the groups in the risk of suicidal ideation, while a relative risk < 1 indicates a protective factor, and a relative risk > 1 indicates a risk factor. In reporting the results, we computed the “change in risk (%)” as the percentual difference from the relative risk value to 1 (1 being the null-value of relative risk), e.g. for a relative risk of 1.4, the change in risk was +40%. The detailed results of all statistical tests are available in the Supplemental material. In the Results chapter, only the values of the significant tests are reported, and otherwise the lack of significance is mentioned.

**Table 1:** Demographic factors influencing suicidal ideation.

| Factor | Comparing variables | With suicidal thoughts | Relative risk of suicidal ideation | Change in risk (%) |
| --- | --- | --- | --- | --- |
| Age | < 33 y. | 6.25% | 0.77<br>for <33 vs. ≥ 33 | - 23% |
|  | ≥ 33 y. | 8.15% |  |  |
| Nr. of children | ≥ 2 | 9% | 1.63<br>for ≥ 2 vs. 1 | + 63% |
|  | 1 | 5.5% |  |  |
| Partnership | No | 8.7% | 1.19<br>for no vs. yes | + 19% |
|  | Yes | 7.3% |  |  |
| Education | Low | 9.1% | 1.3<br>for low vs. high | + 30% |
|  | High | 7.0% |  |  |
| Income | Low | 9.9% | 1.71<br>for low vs. med. | + 71% |
|  | Med. | 5.8% |  |  |
|  | High | 5.7% | 0.98<br>for high vs. med. | - 2% |

□ white = no significant change in suicidality score; grey shades = significant change in suicidality score and an increase in risk between: ■ 1-99%.

## Results

### Demographics

The sample in this study consisted of 1964 mothers, with a mean age of 33.36 years (median = 33 years, range = 18-47 years, SD = 4.3 years). The mothers identified as female (1962, 99.9%) or diverse (2, 0.1%). The children’s age was between 3 and 495 days (mean = 262.4 days, SD = 126.9 days) and the gender was female (948, 47.7%), male (1038, 58.3%), or intersex/not determined (2, 0.1%). For half of the mothers, it was their first child (941, 47.9%), for a third it was the second child (703, 35.8%), and less often the third (251, 12.8%), fourth (48, 2.4%), fifth or further child (21, 1.1%). The pregnancies were usually with one fetus (1941, 98.7%), but also sometimes twins (22, 1.1%) and even triplets (1, 0.05%). Only 1.2% of the participants reported not to be in a relationship (N = 23). The level of education was, on average, high, with 56.9% of mothers having university degrees, 17.2% having finished a professional or technical college or seminary, 19.6% an apprenticeship, 4.7% other form of education and 1.6% with no professional training. The net monthly household income level was reported as low (<2200 Euro) by 36.9% of mothers, medium (2200 – 5800 Euro) by 56%, and high (>5800 Euro) by 7.1%.

### Suicidal ideation: descriptive statistics

While in our sample 1820 mothers (92.7%) stated that they never experienced suicidal thoughts during the seven days before the questionnaire, 144 (7.3%) reported having them with various degrees of frequency. Nine mothers (0.5%) said they often had such thoughts, 50 (2.5%) said sometimes, and 85 (4.3%) said seldom.

### Factors influencing suicidal ideation

#### Demographic factors: age, number of children, partnership, education, income

When comparing younger (<33 years) mothers to older ones (≥33 years) by median split, there was no significant difference between the suicidality score in these two groups, but the younger mothers had a lower risk of suicidal ideation, meaning that the risk was lower in younger mothers, but not enough to reach statistical significance (Table 1; for detailed statistics see Supplemental material). The number of children played a significant role, with mothers having two or more children reporting higher suicidality scores (*W* = 464711, *p =* 0.003), and having a 63% increase in the risk of suicidal ideation compared to mothers with only one child. The presence or absence of a partner did not significantly influence the suicidality score, but the mothers with no partner had a higher risk of suicidal ideation (+19%) compared to those with a partner. That means that the absence of a partner increased the risk of suicidal ideation but not enough to reach statistical significance. The level of education was also not significantly related to the suicidality score, but the risk of suicidal ideation increased 30% if the mothers had lower levels of education (none or apprenticeship) compared to those with higher levels (college and university). This indicates that participants with lower levels of formal education had a higher risk of suicidal ideation than those with higher levels of education, but not enough to reach statistical significance. Lastly, income played a significant role, with lower income levels being related to higher suicidality scores (*W* = 414753, *p* = 0.001) and an increased (+70%) risk of suicidal ideation compared to moderate income levels (Table 1). However, between the participants with a high income and those with a medium income there were no significant differences in the suicidality score or risk of suicidal ideation, indicating that the income is not linearly related to the suicidality risk, but rather reaches a plateau at the medium income level.

#### Social support

During the pandemic, the stay-at-home orders, the quarantines, and the recommended social distancing measures, as well as the reports of higher morbidity from COVID-19 in the elderly, led to a powerful decrease in social contact. To protect themselves, the child, and the grandparents, some families completely cut physical contact with the child’s grandparents (Gulland, 2020). Furthermore, the isolation and added stress of unemployment or simply of the unknown might have increased the levels of conflict between partners and led to a higher rate of domestic violence (Vives-Cases et al., 2021). Therefore, the social support that the mothers had during this time might have changed in many aspects, and that is why, in the current study, we evaluated different forms of social support: (1) a social support score which inquired about the subjective feeling of support, (2) the level of partner support which asked how often one felt well treated by their partner, (3) whether actual physical help was received from the child’s grandparents and (4) whether there was help from other family members and friends. All of these factors strongly influenced the suicidality risk and significantly affected the suicidality score, highlighting the effects of the many forms of social support on suicidal ideation.

##### Social Support Score

A low (below the median) social support score was associated with a significantly higher suicidality score than high (equal or above the median) social support, *W* = 485516, *p <* 0.001, and it increased the relative risk of suicidal ideation by 265% (Table 2).

**Table 2:** Forms of social support and their effect on suicidal ideation.

| Factor | Comparing variables | With suicidal thoughts | Relative risk of suicidal ideation | Change in risk (%) |
| --- | --- | --- | --- | --- |
| **Social Support Score** | Poor | 13.8% | 3.65<br>for poor vs. good | + 265% |
|  | Good | 3.8% |  |  |
| **Well treated by partner** | Occasionally/<br>rarely/never | 17.3% | 2.71<br>for occasionally/rarely/<br>never vs. (almost) always | + 171% |
|  | (Almost) always | 6.4% |  |  |
| **Help from child's grandparents** | No | 10.4% | 1.94<br>for no vs. yes | + 94% |
|  | Yes | 5.4% |  |  |
| **Help from family and friends** | No | 9.0% | 2.28<br>for no vs. yes | + 128% |
|  | Yes | 3.9% |  |  |

□ white = no significant change in suicidality score; grey shades = significant change in suicidality score and an increase in risk between: □ 1-99%; ■ 100-199%; ■ 200-299%.

##### Partner support

The partner support at the time of answering was predominantly good, with over 90% reporting to feel well treated by their partners always or almost always. However, 7.7% only felt well-treated occasionally, and 1% seldom or never. Feeling (almost) always well-treated was associated with significantly lower suicidality scores (*W =* 132568, *p <* 0.001) as compared to feeling occasionally, seldom, or never well-treated. The mothers who occasionally, seldom, or never felt well treated by their partner had a +171% increase in the suicidal ideation risk (Table 2).

##### Help from child’s grandparents

Most parents received help from the grandparents of the child, but almost 40% did not (11% due to the pandemic and 28% due to other reasons). The Wilcoxon rank sum tests showed a significant difference (*W =* 436705, *p <* 0.001) in the suicidality score between the two groups (help and no help), with a higher score for the parents who received no help than those who did. The relative risk ratio of suicidal ideation for the parents who did not receive help increased by 94% compared to those who did (Table 2).

##### Help from family and friends

Only a third (32.4%) of the participants received help from other family members or from their friends, while two thirds did not (25.6% due to the pandemic and 42.1% due to other reasons). The Wilcoxon rank sum test showed a significant difference between the two groups (*W =* 401164, *p <* 0.001), with higher suicidality scores for those who did not receive help. Not receiving help also increased the relative risk ratio of suicidal ideation with 128% (Table 2).

#### Pandemic influence: overall scores and individual questions

##### Pandemic Repercussions Score

The median pandemic repercussions score was 2 (mean = 1.7), on a possible range from -10 (overwhelmingly good influence of the pandemic) to +10 (overwhelmingly poor influence of the pandemic). After a median split, we found that the participants reporting rather negative influences of the pandemic had significantly higher suicidality score than the participants reporting rather positive influences of the pandemic (*W* = 494425, *p <* 0.001). The relative risk of suicidal ideation for the participants who felt more negative influences of the pandemic increased with 163% compared to the participants who felt more positive influences of the pandemic. Consequently, feeling negatively impacted by the pandemic worked as a strong risk factor for suicidal ideation.

The individual components of the pandemic repercussions score weighted differently in their effect on maternal suicidal ideation. The pandemic influence on the **pregnancy**, reported as mostly negative (see Table 3), had no significant effect on the suicidality score, but increased the relative risk of suicidal ideation with 18% in the mothers feeling that the pandemic negatively influenced their pregnancy compared to the mothers who felt that the pandemic had no influence. The relative risk of suicidal ideation in mothers feeling a positive effect of the pandemic on their pregnancy was, in turn, decreased by 15% compared to those feeling no effect. The pandemic influence on the **birth experience** was considered negative in more than half of the participants, while a third reported no influence (Table 3). There was no significant difference in the suicidality score between the participants who felt a bad influence and those who felt no influence, or between those who felt a good influence versus no influence. Both the negative and the positive influence increased the suicidality risk with 41% and 22%, respectively (Table 4). Half of the participants reported that they felt no effect of the pandemic on the **health and development of their child**, while 40% reported a negative impact and 9% reported a positive impact (Table 3). There were significant differences between the groups regarding the suicidality score (negative vs. no effect and positive vs. no effect), with higher scores in those who felt a negative impact and lower scores in those who felt a positive impact, compared to those who felt no effect (*W =* 418176, *p <* 0.001, and *W =* 76806, *p =* 0.001, respectively). The relative risk of suicidal ideation increased with 126% for the participants who felt the pandemic negatively impacted the health and development of their child and decreased with 32% for those who felt a positive impact, compared to those who felt no impact. Consequently, feeling that the pandemic positively influenced the child’s health and development worked as a protective factor, while feeling a negative influence worked as a risk factor for suicidal ideation. Most participants were not affected **financially** by the pandemic, but over a third were negatively impacted and 6% were positively impacted (Table 3). Wilcoxon tests showed a significant difference in the suicidality score between the participants who felt a negative influence on their financial situation and those who felt no influence (*W =* 426778, *p <* 0.001). In the participants whose finances were negatively affected, the relative risk of suicidal ideation increased by 94% compared to those who felt no influence (Table 4). The pandemic influence on the **relationship with the partner** also impacted the suicidality score, so that the participants who felt a negative influence on their partnership had higher scores than those who felt no difference (*W =* 219888, *p <* 0.001) as well as compared to those who felt a positive influence (*W =* 227427, *p <* 0.001). The relative risk of suicidal ideation for the participants whose relationships were negatively affected increased by 223% compared to those who felt no impact. In the participants who felt a positive impact, it decreased by 13%. Consequently, feeling a negative impact of the pandemic on the partnership worked as a risk factor for suicidal ideation, and this again underlines the importance of the relationship with the partner and the social support it provides on the mental health of the mother.

**Table 3:**
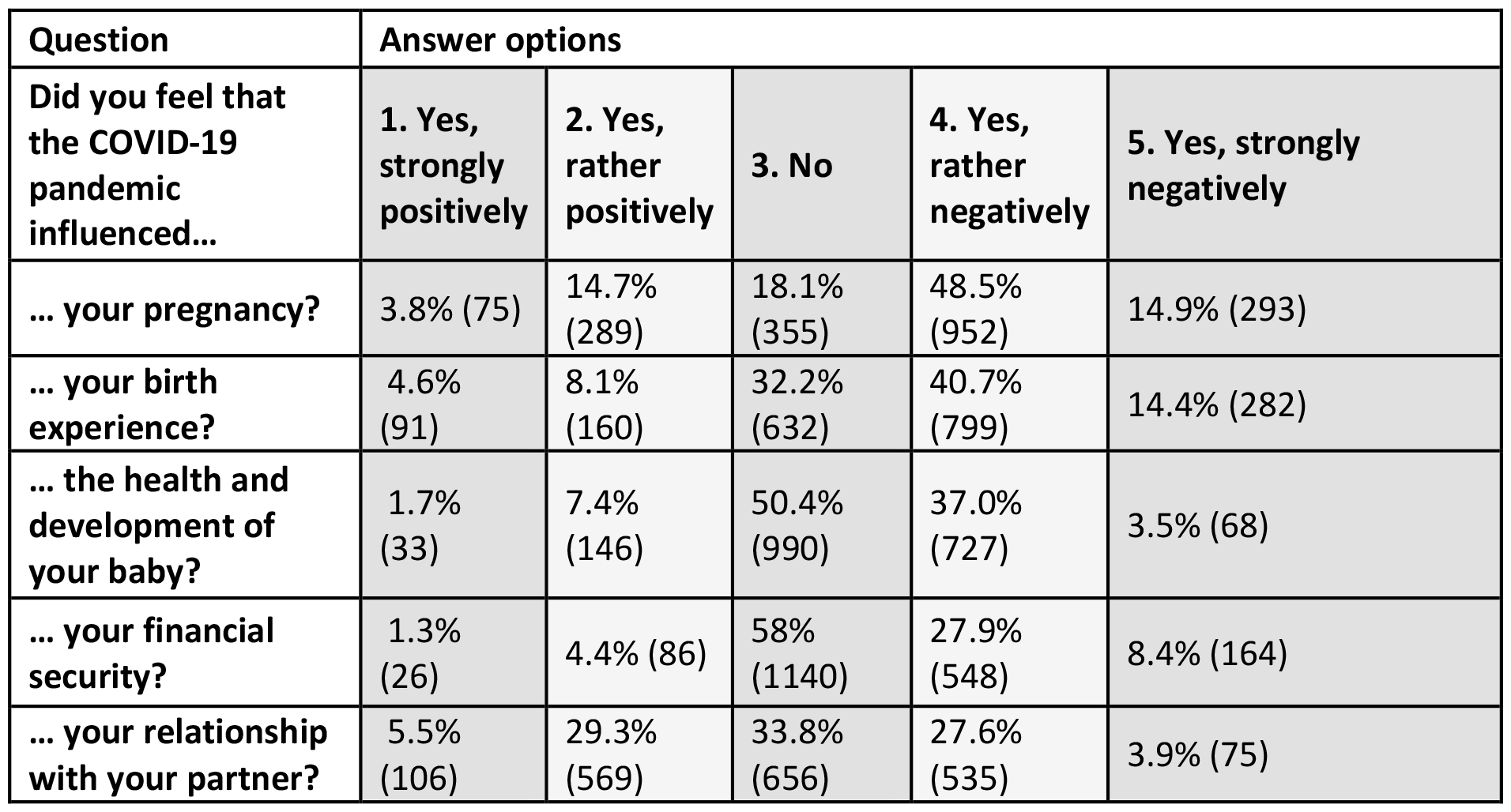
Pandemic Repercussions Score.

| Question | Answer options |  |  |  |  |
| --- | --- | --- | --- | --- | --- |
| Did you feel that the COVID-19 pandemic influenced... | 1. Yes, strongly positively | 2. Yes, rather positively | 3. No | 4. Yes, rather negatively | 5. Yes, strongly negatively |
| ... your pregnancy? | 3.8% (75) | 14.7% (289) | 18.1% (355) | 48.5% (952) | 14.9% (293) |
| ... your birth experience? | 4.6% (91) | 8.1% (160) | 32.2% (632) | 40.7% (799) | 14.4% (282) |
| ... the health and development of your baby? | 1.7% (33) | 7.4% (146) | 50.4% (990) | 37.0% (727) | 3.5% (68) |
| ... your financial security? | 1.3% (26) | 4.4% (86) | 58% (1140) | 27.9% (548) | 8.4% (164) |
| ... your relationship with your partner? | 5.5% (106) | 29.3% (569) | 33.8% (656) | 27.6% (535) | 3.9% (75) |

**Table 4:** The effects of the pandemic on different aspects of life and their consequences on suicidal ideation.

| Effect of the pandemic on... | The influence was... | With suicidal thoughts | Relative risk of suicidal ideation | Change in risk (%) |
| --- | --- | --- | --- | --- |
| ... the pregnancy | Negative | 8.0% | 1.18<br>for negative vs. no infl. | +18% |
|  | No influence | 6.8% |  |  |
|  | Positive | 5.8% | 0.85<br>for positive vs. no influence | -15% |
| ... the birth experience | Negative | 8.2% | 1.41<br>for negative vs. no infl. | + 41% |
|  | No influence | 5.9% |  |  |
|  | Positive | 7.2% | 1.22<br>for positive vs. no influence | + 22% |
| ... the health and development of the child | Negative | 11.9% | 2.26<br>for negative vs. no infl. | + 126% |
|  | No influence | 4.9% |  |  |
|  | Positive | 3.4% | 0.68<br>for positive vs. no influence | -32% |
| ... the financial stability | Negative | 10.5% | 1.94<br>for negative vs. no infl. | +94% |
|  | No influence | 5.4% |  |  |
|  | Positive | 6.3% | 1.15<br>for positive vs. no influence | +15% |
| ... the relationship with the partner | Negative | 14.3% | 3.23<br>for negative vs. no infl. | +223% |
|  | No influence | 4.4% |  |  |
|  | Positive | 3.9% | 0.87<br>for positive vs. no influence | -13% |

□ white = no significant change in suicidality score; grey shades = significant change in suicidality score and an increase in risk between: □ 1-99%; ■ 100-199%; ■ 200-299%.

Overall, the negative effects of the pandemic that contributed most to the increase in suicidal risk were those that the mothers perceived on the relationship with their partner, on the health and development of the child, and on the financial stability of the family (Table 4).

##### Pandemic Distress Score

The median pandemic distress score was 5 (mean = 5.12) on a possible range from 0 (no distress) to 12 (high distress). The participants with a score higher than 5 (high levels of pandemic distress) had significantly lower suicidality score than the participants with a score of 5 or lower (*W =* 392738, *p =* 0.006). Surprisingly, worrying more about the disease decreased the relative risk of suicidal ideation by 39%.

The components of the pandemic distress score consisted in the worry about someone close, oneself, or the baby getting sick with COVID-19 (Table 5), and each had a different effect on the suicidality score and on the suicidal ideation risk (Table 6). Worrying that **people important to oneself** would get sick of COVID-19 made a significant difference in the suicidality score, so that the participants who worried slightly had a higher suicidality score than those who worried moderately (*W =* 235353, *p <* 0.001). However, these two groups did not differ from those who worry strongly. Compared to moderate worry, strong worry increased the risk of suicidal ideation by 48% while slight worry increased it by 112%. Worrying that **oneself** will get sick also played a significant role, with participants who worried slightly having higher suicidality scores than those who worried moderately (*W =* 289404, *p =* 0.01). The mean suicidality score for those who worried strongly was not significantly different from the other two groups. However, both worrying slightly and worrying strongly about one’s health increased the risk ratio for suicidal ideation (compared to moderate worry) with 65% and 20%, respectively. Worrying about **the baby** did not significantly change the suicidality scores. The risk ratio of suicidal ideation for worrying strongly vs. moderately was increased with just 5%, and for worrying slightly vs. moderately with 39%.

**Table 5:** Pandemic Distress Score.

| Table 5: Pandemic Distress Score |  |  |  |  |  |
| --- | --- | --- | --- | --- | --- |
| Question | Answer options |  |  |  |  |
| Do you worry that a COVID-19 infection... | 1. Very slightly | 2. Slightly | 3. Moderately | 4. Strongly | 5. Very strongly |
| ... could affect someone important to you? | 17.6% (346) | 17.3% (339) | 33.2% (652) | 21.7% (427) | 10.2% (200) |
| ... could affect you? | 28.0% (505) | 25.7% (464) | 32.0% (578) | 10.3% (185) | 4% (72) |
| ... could harm your baby? | 22.2% (436) | 20.3% (399) | 26.2% (515) | 18.6% (365) | 12.7% (249) |

**Table 6:**
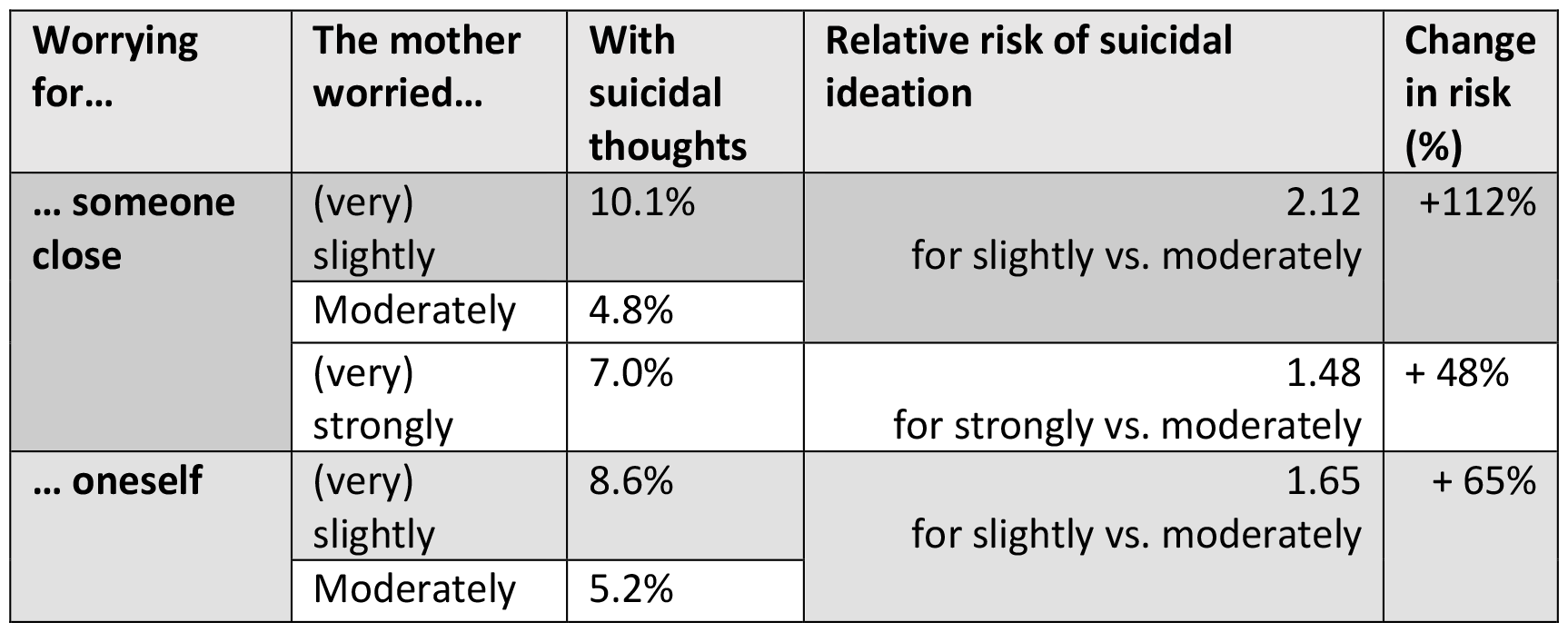
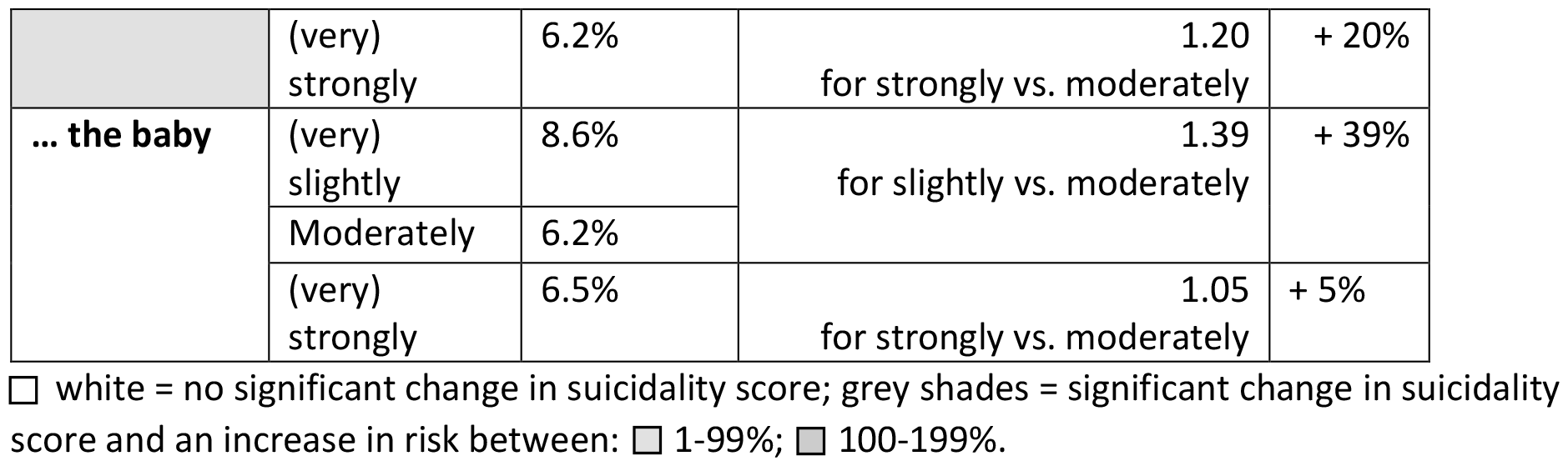
Sources of pandemic distress and their effect on suicidal ideation.

| Worrying for... | The mother worried... | With suicidal thoughts | Relative risk of suicidal ideation | Change in risk (%) |
| --- | --- | --- | --- | --- |
| ... someone close | (very) slightly | 10.1% | 2.12<br>for slightly vs. moderately | +112% |
|  | Moderately | 4.8% |  |  |
|  | (very) strongly | 7.0% | 1.48<br>for strongly vs. moderately | + 48% |
| ... oneself | (very) slightly | 8.6% | 1.65<br>for slightly vs. moderately | + 65% |
|  | Moderately | 5.2% |  |  |
|  | (very) strongly | 6.2% | 1.20<br>for strongly vs. moderately | + 20% |
| ... the baby | (very) slightly | 8.6% | 1.39<br>for slightly vs. moderately | + 39% |
|  | Moderately | 6.2% |  |  |
|  | (very) strongly | 6.5% | 1.05<br>for strongly vs. moderately | + 5% |

□ white = no significant change in suicidality score; grey shades = significant change in suicidality score and an increase in risk between: □ 1-99%; ■ 100-199%.

These results indicate a non-linear correlation between the level of worry and the suicidal risk, with a higher risk at the extremes and lower risk in the middle.

#### Stress levels

Stress levels were assessed using the PSS-10 score. The median score was 18 (mean = 17.7, possible range 0-40), with higher scores indicating more stress (0-13: low stress, 14-26: moderate stress, 27-40: high stress). Two thirds of the participants (63%) were moderately stressed, a quarter had low stress levels (27.8%) and 9.2% were highly stressed. Wilcoxon tests revealed highly significant differences between the three groups. The highly stressed group had a much higher suicidality score than the moderately-stressed (*W =* 139437, *p <* 0.001) and than the low-stressed (*W =* 64435, *p <* 0.001) groups. The low-stressed group had a much lower suicidality score than the moderately stressed group (*W =* 31635, *p <* 0.001). Compared to a moderate stress level, a high stress level increased the risk ratio for suicidal ideation with 350%, while the lower stress levels decreased it with 82%. These results highlight the stress levels as the strongest risk factor for suicidal ideation.

#### Access to a midwife, quarantine and coping mechanisms

Over half of the participants reported that the COVID-19 pandemic led to no restriction in their access to a midwife. However, over a quarter felt their access was slightly restricted and a fifth moderately or strongly restricted (Table 7). A Wilcoxon test between the participants with good access (not or slightly restricted) and those with poor access (moderately or strongly restricted) revealed no significant differences in their suicidality scores, though a trend (*W =* 313844, *p =* 0.08) towards a higher suicidality score for those with poor access. The risk ratio for suicidal ideation in those with poor access was increased by 38%.

**Table 7:** Access to a midwife, quarantine and coping mechanisms.

| Question | Answer | % (N) | Groups and % of suicidal ideation | Change in risk (%) |
| --- | --- | --- | --- | --- |
| Was the access to a midwife restricted during the pregnancy or after birth restricted due to COVID-19? | No, not really | 50.7% (996) | Good access: 6.8% | + 38%<br>for poor vs. good access |
|  | Yes, slightly | 28.1% (552) |  |  |
|  | Yes, moderately | 12.0% (235) | Poor access: 9.4% |  |
|  | Yes, strongly | 9.2% (181) |  |  |
| Have you been in quarantine during the pregnancy, birth or postpartum? | Yes | 20.2% (397) | Quarantine: 6.8% | -9%<br>for having been in quarantine vs. not having been in quarantine |
|  | No | 79.8% (1567) | No quarantine: 7.5% |  |
| Is there something that helps you during the Corona-situation? (E.g., hobbies, sport, religion, etc.) | Yes, and it helps me a lot | 41.6% (817) | Effective coping mechanism: 5.8% | + 132%<br>for not having an effective coping mechanism vs. having one |
|  | Yes, and it helps me moderately | 37.9% (744) |  |  |
|  | Yes, but it only helps a little | 8.2% (161) | No effective coping mechanism: 13.4% |  |
|  | No, nothing actually helps | 12.3% (242) |  |  |

□ white = no significant change in suicidality score; grey shades = significant change in suicidality score and an increase in risk between: □ 1-99%; ■ 100-199%.

A fifth of our sample had been in quarantine before, during or after birth (Table 7). The quarantine did not significantly influence the suicidality score or the risk of suicidal ideation (Table 7). The mothers were asked whether they have a coping mechanism that helps them deal with the pandemic situation. Most of them answered positively, but 20% found little or no help in a coping mechanism. The coping mechanism had a significant impact in the suicidality score (*W =* 290587, *p <* 0.001), with a higher suicidality score for the mothers without an effective coping mechanism than the mothers who had one. The relative risk ratio of suicidal ideation for those not having an effective coping mechanism compared to those having one was increased by 132%.

#### Baby health: type of birth, birth weight, gestational age at birth

The type of birth (26% caesarean, 7% suction or forceps, 66% natural) had no significant association with the suicidality score, and neither did the baby’s birth weight (4.7% low < 2500g, 83% normal 2500-400g, 2% high > 4000g) or gestational age at birth (7.6% preterm, 85.9% at term, 6.5% post-term) (Table S1, Supplemental Material).

## Discussion

The prevalence of suicidal ideation of 7.3% in postpartum mothers corresponds to a global average over 71 studies from 23 countries published in a previous meta-analysis (Xiao et al., 2022). However, compared to a study on 306 German mothers (Martini et al., 2019), which was performed during 2009-2010 and reported a prevalence of 3.5%, our sample’s prevalence was twice as high. Furthermore, the sample from Martini et al. (2019) and our sample are more comparable because participants in both studies have higher levels of education and are more likely to be in a relationship than the general population (Florea et al., 2023). The increase in the prevalence of suicidal ideation in the German-speaking area might be related to a general worsening of mental health during the COVID-19 pandemic as reported by others (Beutel et al., 2021; Pan et al., 2021; Vindegaard & Benros, 2020; Zanardo et al., 2020).

The analysis of the relative risk of suicidal ideation indicated that the strongest risk factors (relative risk > 3) were high stress levels, a lack of social support, and feeling that the pandemic had a bad influence on the relationship with one’s partner. Furthermore, moderate risk factors (relative risk > 2) were perceiving a negative effect of the pandemic on the health and development of the child, not worrying about other people getting sick of COVID-19, feeling poorly supported by one’s partner, receiving no help from family members or friends, and lacking an effective coping mechanism. Weaker risk factors (relative risk > 1) were having multiple children, having a lower income, feeling that the pandemic had a bad influence on the birth experience and on the financial security, not worrying that oneself would get sick, having restricted access to a midwife, and receiving no help from the child’s grandparents. Protective factors (relative risk < 1) were a high pandemic distress score and (especially) a low stress level.

Considering these results, our hypothesis that social support has a protective influence against suicidal ideation was confirmed. The same can be said about the hypothesis that the perceived negative effects of the pandemic would correlate with a higher risk of suicidal ideation. Furthermore, the hypothesised risk factors of high stress levels, lower income, and poor access to a midwife were also confirmed. The hypothesis that a lower education level would also be correlated with a higher suicidal ideation was partly true, as the relative risk was 1.3 for low education vs. high education levels, indicating that a lower education level is a weak risk factor, but the Wilcoxon test of suicidality score between the two groups was not significant.

These results are consistent with findings from previous studies, which have identified correlations between income (Begum et al., 2021), a lack of support from one’s partner (Orsolini et al., 2016), and a higher number of children (Howard et al., 2011) with suicidal ideation. Furthermore, the protective effect of social support has also been widely reported (Park et al., 2010; see Reid et al., 2022 for a review). The presence of an effective coping mechanism such as sport, religion or a hobby might reduce the suicidal ideation through distracting from repetitive thoughts, releasing stress, or believing in a higher purpose. The protective effect of such a coping mechanism can be found in the literature, since having little time for hobbies and giving low importance to religion has been previously associated with higher risk of suicidal ideation (Lamlé et al., 2023), while regular physical activity has been found to prevent stress and suicidal ideation (Brailovskaia et al., 2023; Koo & Kim, 2020).

At first glance it may seem counterintuitive that not worrying about other people or oneself getting sick of COVID-19 poses a risk factor for suicidal ideation. Yet, not worrying about the virus did not exclude other sources of distress. Businesses suffered, many people lost their jobs or had to take unpaid leave of absence, which threatened their financial security. Border crossing became strongly regulated in places where until then there had been no restrictions (Schengen space), affecting especially families split across borders. Those who did not see the virus as a significant threat to their or others’ lives, might have considered these and other restrictions unwarranted and as a potential attack to “fundamental rights and freedom of expression”. Such perceptions likely subjected them to significant social pressure and stress, contributing to suicidal thoughts (Schabus et al., 2022). Another possibility might be that the segment of population that did not worry about the pandemic felt marginalized by the rest of the society and forced to submit to protective measures that they did not consider necessary or appropriate (Hannawa & Stojanov, 2024; Jaspal & Nerlich, 2023). Indeed, our data shows that the worry about others, oneself, or the child getting ill with COVID-19 was strongly and positively correlated with one’s perception on COVID-19 severity and with one’s agreement with the authorities on the imposed restrictions (rho values around 0.5, see Suplemental material for detailed statistics). Having to act contrary to their beliefs might have led to frustration, while criticising the measures might have put them in conflict with others, leading to more isolation and an environment of hostility. All these effects might increase the depressive symptoms and the suicidality ideation. However, this comes in contrast to other studies, who found those who perceived less risk from the new coronavirus to have lower depression levels (Duffy & Allington, 2020; Hannawa & Stojanov, 2024) or psychological distress (Schneider et al., 2021) than those who perceived the threat as very high but did not trust the government to impose the right measures. However, our study and the ones by Duffy and Hannawa are consistent in finding that the lowest levels of depression were in the “moderate worry” group, i.e. those participants who perceived the virus as moderately dangerous.

Through exploration, this study further found that the relative risk of suicidal ideation is increased for the mothers who receive no help from their family or friends, which is a practical side of social support, apart from the emotional, perceived social support, as assessed by our social support questions. The perceived negative effects of the pandemic on the health and development of the child, on the financial security and on the delivery experience were further risk factors of suicidal ideation, and they fall in line with the correlation between perceived negative pandemic effects and depression scores found in the previous study (Florea et al., 2023). These perceived negative effects might lead to a feeling of helplessness, increasing the suicidality risk. Similarly, a pandemic-imposed perceived restricted access to a midwife might raise the mother’s insecurity about childrearing and lead to the same feeling of failure and low self-esteem associated with depression and suicidal ideation.

Overall, the current analysis of suicidal ideation in Austrian and German postpartum mothers points towards an increased prevalence compared to previous reports, and highlights the importance of social support in all its forms, and of reducing stress levels in different aspects of life (general stress, financial stress, child development-related stress). These risk and protective factors could be targets of social and public health policies, while the first step should be a general screening program for depression and suicidality in postpartum women.

## Limitations

One limitation is the possible self-recruiting bias of the online study. Advertised on parents’ social media groups and on the radio, the survey was open to anyone meeting the criteria, and the sample was therefore self-selected to those participants interested in the study. Another limitation is the differences between our sample and the general population (e.g., level of education, prevalence of single mothers; for more details see Florea et al. (2023), *Methods*), which might reduce the generalizability of the results. A further limitation is that the used item of the EPDS asks about self-harm and not explicitly about thoughts of suicide, but has been widely used as a proxy for suicidal ideation (Martini et al., 2019; Xiao et al., 2022).

## Conclusion

The current study indicates that suicidal ideation is more frequent in German and Austrian postpartum mothers than previously thought, reaching double the values found 15 years ago (Martini et al., 2019). We reveal that the main risk is stress, and the main protective factors are social and partner support. Without doubt suicidal ideation can be seen as a public health problem, and maternal suicidality should be screened more broadly, especially during times of crisis and heightened stress in the society. As a countermeasure, we encourage to better communicate the importance of strong social-support networks for mothers at risk, and to provide better and easier access to stress reduction and stress resilience trainings for young families.

## Data Availability

All data produced in the present study are available upon reasonable request to the authors.

