## Supplemental Material for "Postpartum suicidal ideation in Austria and Germany during the COVID-19 pandemic"

**Table S1: Summary of results**

| **Suicidality score results and relative risk ratios for suicidal ideation** | | | |
|---|---|---|---|
| **Factor** | **Comparing variables** | **Suicidality score - Statistical test significance** | **Change in risk (%)** |
| **Age** | <33 vs. ≥ 33 | $W$ = 481952, $p$ = 0.119 | -23% |
| **Nr children** | ≥ 2 vs. 1 | $W$ = 464711, $p$ = 0.003 | +63% |
| **Partnership** | no vs. yes | $W$ = 22005, $p$ = 0.796 | +19% |
| **Education** | 0 vs. 1 vs. 2 vs. 3 | $\chi^2$ = 4.786, df = 3, $p$ = 0.188 | |
| | low (0 or 1) vs. high (2 or 3) | $W$ = 309742, $p$ = 0.151 | +30% |
| **Income** | low vs. med. vs. high | $\chi^2$ = 6.705, $p$ = 0.082 | |
| | low vs. med. | $W$ = 414753, $p$ = 0.001 | +71% |
| | high vs. med. | $W$ = 76831, $p$ = 0.951 | -2% |
| **Social support** | poor vs. good | $W$ = 180082, $p$ < 0.001 | +265% |
| **Pandemic Repercussions Score (PRS)** | >2 (neg. influence) vs. ≤ 2 (pos. influence) | $W$ = 494425, $p$ < 0.001 | +163% |
| **PRS - Pregnancy** | bad vs. no influence | $W$ = 223595, $p$ = 0.462 | +18% |
| | good vs. no influence | $W$ = 63969, $p$ = 0.583 | -14% |
| **PRS - Birth** | bad vs. no influence | $W$ = 349682, $p$ = 0.07 | +41% |
| | good vs. no influence | $W$ = 80315, $p$ = 0.486 | +22% |
| **PRS - Health and dev. Child** | bad vs. no influence | $W$ = 418176, $p$ < 0.001 | +126% |
| | good vs. no influence | $W$ = 87153, $p$ = 0.341 | -32% |
| **PRS - Financial security** | bad vs. no influence | $W$ = 426778, $p$ < 0.001 | +94% |
| | good vs. no influence | $W$ = 64400, $p$ = 0.698 | +15% |
| **PRS - Partnership** | bad vs. no influence | $W$ = 219888, $p$ < 0.001 | +223% |
| | good vs. no influence | $W$ = 220134, $p$ = 0.601 | -13% |
| **Pandemic Distress Score (PDS)** | high (> 5) vs. low (≤ 5) | $W$ = 392738, $p$ = 0.006 | -39% |
| **PDS - Close people sick** | slight vs. moderate worry | $W$ = 235353, $p$ < 0.001 | +112% |
| | strong vs. moderate worry | $W$ = 209236, $p$ = 0.072 | +48% |
| **PDS - Self sick** | slight vs. moderate worry | $W$ = 289404, $p$ = 0.015 | +65% |
| | strong vs. moderate worry | $W$ = 75046, $p$ = 0.544 | +20% |
| **PDS - COVID-19 baby** | slight vs. moderate worry | $W$ = 159154, $p$ = 0.806 | +39% |
| | strong vs. moderate worry | $W$ = 150540, $p$ = 0.439 | +5% |

| | | | |
|---|---|---|---|
| **Stress (PSS)** | high vs. moderate | $W$ = 139437, $p$ < 0.001 | +350% |
| | low vs. moderate | $W$ = 316365, $p$ < 0.001 | -92% |
| **Access to a midwife** | moderately/strongly restricted vs. not/slightly restricted | $W$ = 313822, $p$ = 0.079 | +38% |
| **Birth type** | cesarean vs. natural | $W$ = 342461, $p$ = 0.407 | +16% |
| **Birth weight** | low vs. normal vs. high | $\chi^2$ = 0.81, df = 3, $p$ = 0.847 | |
| | low vs. normal | $W$ = 65192, $p$ = 0.938 | -4% |
| | high vs. normal | $W$ = 150028, $p$ = 0.843 | +5% |
| **Gestational age** | preterm vs. term vs. postterm | $\chi^2$ = 0.89, df = 3, $p$ = 0.829 | |
| | preterm vs. term | $W$ = 123222, $p$ = 0.379 | -28% |
| | postterm vs. term | $W$ = 108266, $p$ = 0.909 | +5% |
| **Quarantine** | yes vs. no | $W$ = 309052, $p$ = 0.661 | -9% |
| **Partner support** | poor vs. good | $W$ = 132568, $p$ < 0.001 | +171% |
| **Help from child's grandparents** | no vs. yes | $W$ = 436705, $p$ < 0.001 | +94% |
| **Help from family and friends** | no vs. yes | $W$ = 401164, $p$ < 0.001 | +128% |
| **Effective coping mechanism** | no vs. yes | $W$ = 290587, $p$ < 0.001 | +132% |

□ white = no significant change in suicidality score but may still represent a risk or protective factor as shown by the risk change); □ yellow = significant change in suicidality score and an increase in risk between 1-99%; □ orange = significant change in suicidality score and an increase in risk between 100-199%; □ red = significant change in suicidality score and an increase in risk between 200-299%; purple □ = significant change in suicidality score and an increase in risk > 300%; □ Green = protective factor. In the case of trends (p < 0.1), the row is left white, and the p value is written down.

**Correlations between worry that oneself/family and friends/the child would get COVID-19 and the agreement with the authorities on the restrictions and on COVID-19 severity**

Spearman correlations:

| | Worry about others getting sick | Worry about oneself getting sick | Worry about baby getting sick |
|---|---|---|---|
| Perception of COVID-19 severity | $p$ < 0.001, rho = 0.57 | $p$ < 0.001, rho = 0.53 | $p$ < 0.001, rho = 0.48 |
| Agreement with restrictions | $p$ < 0.001, rho = 0.55 | $p$ < 0.001, rho = 0.51 | $p$ < 0.001, rho = 0.49 |

A moderation analysis using a linear model to test if one's opinion on the restrictions would moderate the relationship between worry for self/others/baby getting sick and the suicidal ideation was not possible. That is because the relationship between the level of worry and the suicidal ideation was not linear, with the moderate levels of worry having the lowest scores of suicidal ideation, while not worrying at all or worrying very much had higher scores of suicidal ideation.
